# Investigating the origin of the Belgian second SARS-CoV-2 wave by using (pre)admission screening samples

**DOI:** 10.1101/2020.12.09.20246462

**Authors:** Reinout Naesens, Laura Heireman, Sarah Vandamme, Philippe Willems, Bruno Van Herendael, Walter Verstrepen, Pieter De Schouwer, Peggy Bruynseels

## Abstract

The goal of this study was to estimate rates of SARS-CoV-2 carriership and viral loads in the general Antwerp population and to compare the estimated prevalences and incidences with governmental data (numbers of detected positive cases, stringency measure index) in order to evaluate the dynamics leading to the second wave. We used (pre)admission screening results from the major Antwerp hospitals for estimating community prevalences and incidences. 43.545 samples were included (April – November 2020). High SARS-CoV-2 carriership rates (mean week prevalence of 1.3%) were found in the general Antwerp population. 35.4% of positive cases carried high viral loads. Only a small proportion (15.3%) of the viral circulation was detected by the nationally implemented testing policy. In the weeks before the second Belgian wave, increasing prevalences and incidences were found, together with country-wide easing of restriction measures. In our opinion these findings have led to origin of the second viral wave.

## INTRODUCTION

Since March 2020, the world has been in the grips of a pandemic caused by the Severe Acute Respiratory Syndrome virus type 2 (SARS-CoV-2) leading to excess mortality and morbidity. The virus has a detrimental impact on both economic and psycho-social well-being^1^.

Belgium was highly impacted, having had to deal with two waves up to November 2020: the first from the beginning of March to the end of May, the second during the fall (showing a decline in November). The first wave was contained by implementing a full lockdown starting on 14 March 2020. The first measures to ease restrictions were introduced on 4 June 2020.

Despite warnings from various Belgian key expert virologists and epidemiologists in September 2020, the government decided to ease restrictions further, having been encouraged by various pressure groups and public opinion. Belgium faced its second wave as from October 2020 (Figure 1).

**Figure 1.**
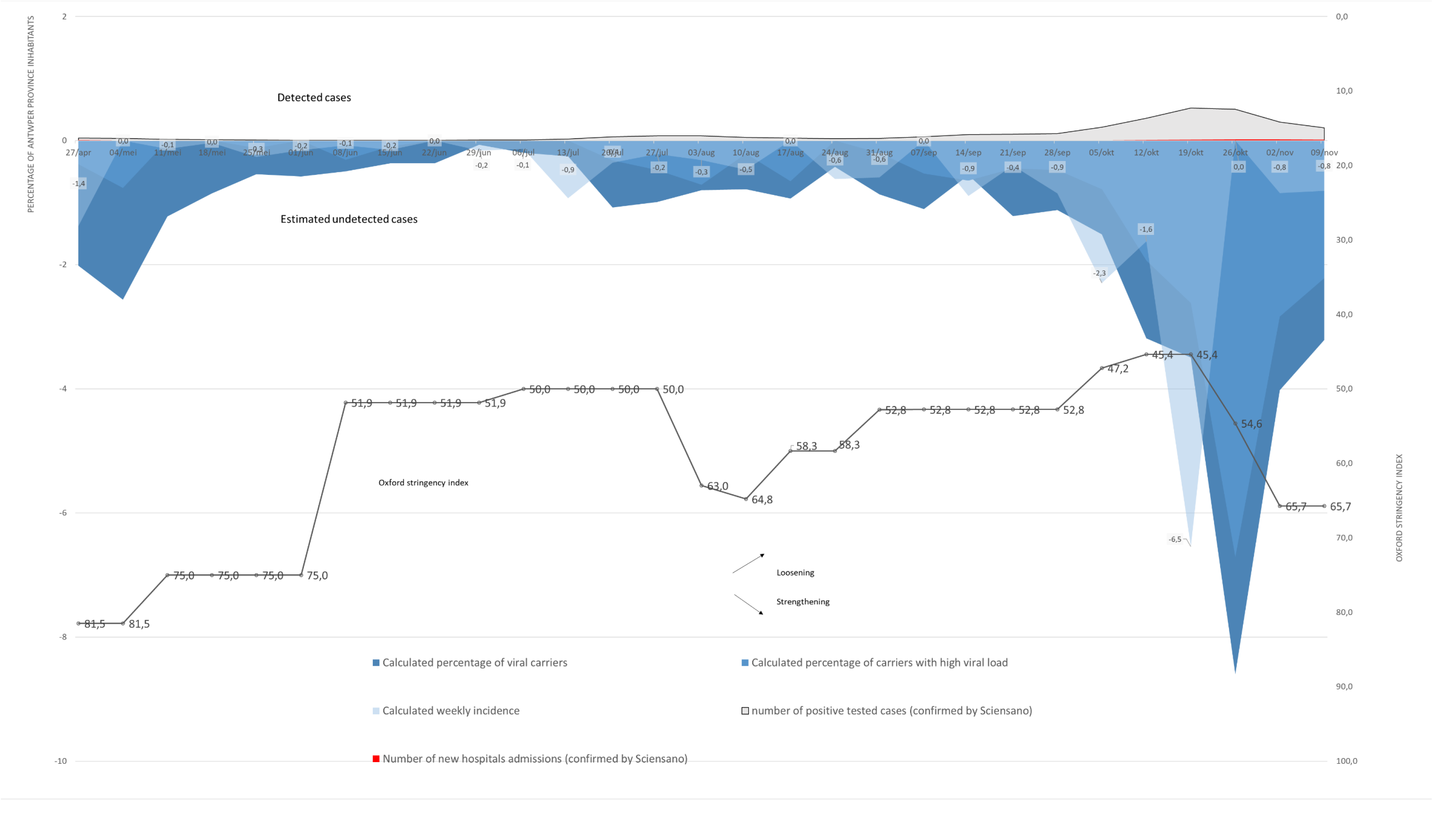
Estimated prevalence and incidence of SARS-CoV-2 carriers, estimated prevalence of carriers with high viral loads, percentages of cases with confirmed a positive SARS-CoV-2 test and percentages of confirmed hospital admitted COVID cases in the Antwerp province.

Until now, little is known about the dynamics and prevalence of asymptomatic carriership and viral loads in the general Belgian population^2^. Furthermore, the role of the different age categories in the viral spread has not been fully elucidated^3^.

Since the major Antwerp hospitals implemented extensive (pre-)admission screening of asymptomatic cases in April 2020 (eg. prior to elective surgery or endoscopic procedures), and have continued to do so, they possess valuable information regarding the actual prevalence of the virus in the general population throughout the pandemic period.

The goal of this study was to determine rates of carriership and viral loads in the general population and to evaluate the dynamics leading to the onset of the second Belgian wave.

## METHODS

### DATA COLLECTION

During the period 27 April to 15 November 2020, we collected SARS-CoV-2 positivity data from (pre)admission screenings from the following hospitals: the Antwerp University Hospital (UZA), the multiple site public hospital ZiekenhuisNetwerk Antwerpen (ZNA), and the major multiple site private Antwerp hospital GemeenschapsZusters Antwerpen (GZA). Together they account for approximately 90% of hospital bed capacity (3697 hospital beds) in the city of Antwerp.

(Pre-)admission screenings were defined as follows: pre-hospital screenings prior to elective admissions (GZA, UZA, ZNA), and systematic admission screenings in asymptomatic patients admitted for other reasons than COVID-19 (GZA, UZA). Screenings of high/low risk contacts or individuals with symptoms were excluded.

A distinction in test indications was obtained by implementing different sample flows per indication (pre-admission screenings were performed by a nurse at patients’ homes for the inclusions from UZA and ZNA, and screenings performed in a drive-through swab laboratory, or during a pre-visit at the swab laboratory, from UZA and GZA respectively). Unique test codes were also used by GZA and UZA for registering the swab samples in the Laboratory Information System (LIS).

Samples were analysed in the laboratory attached to the hospital of the prescribing physician. All laboratories are certified by the Belgian government (BELAC). The following molecular techniques and platforms were used:

In the clinical laboratory of GZA, nucleic acids were extracted on STARlet IVD® (Seegene INC., Seoul, Korea) using the Viral DNA/RNA C-kit. Subsequently SARS-CoV-2 qPCR (CDC N1 target) was performed on a Quantstudio 7 flex qPCR cycler® of Thermo Fisher Scientific (Waltham, MA, USA) according to the protocol published by the Centers of Disease Control and Prevention (CDC).

The laboratory of UZA used the Xpert Xpress SARS-CoV-2 test detecting E and N2 target on the GeneXpert® Platform (Cepheid, USA) as per manufacturer’s instructions. Alternatively, an in-house PCR was performed detecting E target according to Corman *et al*. 2020 with extraction on NucliSens EasyMag® (Biomérieux, France) or QiaSymphony® (Qiagen, The Netherlands) and amplification with Cobas LightCycler 480 II® or Cobas z480 (Roche Molecular Diagnostics, Switzerland). Alternatively, a BD MAX SARS-CoV-2 kit was used detecting N1 and N2 target with an all-in one extraction amplification on the BD MAX® platform (Becton Dickinson, USA).

In the laboratory of ZNA, nucleic acids were extracted on MagNA Pure 96 using the MagNA Pure 96 DNA and Viral NA Small Volume Kit (Roche Molecular Diagnostics, Switzerland). Subsequently SARS-CoV-2 qPCR (N1 target) was performed on a LightCycler 480 qPCR cycler according to the protocol published by the CDC. Alternatively, nucleic acids were extracted on KingFisher Flex using the MagMAX Viral/Pathogen II Nucleic Acid Isolation Kit. These extracts were analysed on a Quantstudio 5 qPCR cycler using the TaqPath™ COVID-19 CE-IVD RT-PCR Kit (ThermoFisher Scientific).

The parameters postal code, age, PCR result (associated with PCR platform used) and cycle threshold (Ct) value (or alternatively Crossing point (Cp)) if available (GZA, ZNA) were retrieved from the LIS for each patient, and transferred to a central ZNA-based and secured database.

The following data were collected from the Belgian governmental scientific institution, Sciensano^4^: absolute numbers (weekly) of positively detected cases for Antwerp province, admitted patients at the hospital for Antwerp province, and returning travelers in August 2020 with positivity rate (only available on national level).

The following data were collected from the Flemish Agency for Care and Health (governmental agency responsible for the contact-tracing policy) for Antwerp province by using the information from the web-based and secured database (Controle-toren)^5^: percentages (weekly) of successfully (by phone) contacted positive cases in the Antwerp province. Age distributions for the Antwerp province were obtained from Statistiek Vlaanderen^6^.

We used the Oxford COVID-19 Government Response Stringency index to estimate governmental measures. The index goes from zero (no restrictions) to hundred (strictest)^7^.

### DATA ANALYSES AND DEFINITIONS

Cases with domicile outside the Antwerp province were excluded for further analysis. SARS-CoV-2 carriership was estimated by calculation of the weekly asymptomatic positivity rate in the study population. The incidence was calculated by using an estimated duration of PCR positiveness of seventeen days as was found by Cevik and colleagues^8^. The percentage of the cohort infected by SARS-CoV-2 (for the whole investigated period) was calculated by adding up all estimated week incidences. By dividing the absolute weekly numbers of positively tested cases in the province by the number of province inhabitants (1.858 million) in the province, the percentage of positively tested patients in the Antwerp province was calculated. The percentage of weekly new COVID-19 hospital admitted patients in the Antwerp province was calculated by dividing the absolute weekly number of admitted patients by the number of inhabitants in the province.

Viral loads (only available for GZA and ZNA) were calculated by extrapolation of the Ct/Cp values corresponding to 5000 genome equivalents per mL of the AccuPlex™ SARS-CoV-2 full genome reference material (Seracare). A high viral load was defined as >= 256 000 viral copies/mL^9^. Ratios of positive cases (on a weekly basis; all inclusions) with high viral loads were plotted on the same timeline as the plotted estimated incidences.

Mean positivity ratios per age decade (0-10 y, 11-20 y, 21-30 y, 31-40 y, 41-50 y, 51-60 y, 61-70 y, 71-80 y, 81-90 y, 90+ y) were investigated for the investigated cohort. Positivity ratios of carriership were calculated for preschools (0-4 y), primary schools (5-11 y) and secondary schools (12-18 y). The mean positivity ratio was also assessed for (pre)school children (0-18 y) and the elderly (+80 y). Box-whisker plots represent the viral load among different age categories.

Ethical approvals were obtained by the hospital Institutional Boards (GZA: Approval N° 200906RETRO; UZA: Approval N°001355; ZNA: Approval N° 5416).

## RESULTS

Overall, 43.545 cases were identified of which 38.763 cases (89.0%) were included. The results of 4782 cases (11.0%) were excluded since domicile (based on postal code) was outside Antwerp province. GZA, UZA, and ZNA accounted for respectively 53.2%, 34.6% and 10.7% of included cases. The overall SARS-CoV-2 positivity rate was 1.3% (n=520). Younger age groups (<21 years old) were underrepresented in the investigated cohort as compared to the Antwerp province (9.0 % versus 22.3%; *p* <0.01). Age groups of > 70 years old were overrepresented in the cohort (25.0 % versus 14.1 %, *p*<0.01). The distribution of cases was not equally dispersed throughout Antwerp province with an underweight of cases from the city of Antwerp versus the Antwerp province (20.8% of cases had domicile in the city of Antwerp, versus 26.8 % of inhabitants for the whole Antwerp province *p* <0.01). Cases with domicile in the city of Antwerp (n = 8067) had an overall positivity rate of 1.8%, whereas cases with domicile outside the city of Antwerp (n = 30696) had an overall positivity rate of 1.2% (*p* < 0.01).

Estimated carriership (with high viral loads), estimated weekly new cases and number of admissions are shown in Figure 1. Calculated weekly cohort carriership varied between 0.1 % (week of 29 June) and 8.6 % (week of 26 October). The mean week percentage was 1.5%. The calculated ratio of carriership with high viral loads varied between 0% (week of 11 May, 18 May, 1 June, 22 June, 29 June, 6 July, 13 July and 24 August) and 6.7 % (week of 26 October). The mean week ratio was 0.8%. Overall, the percentage of positive carriers with high viral loads was 35.4 %. The calculated weekly incidence varied between 0.0% (see Figure 2) and 6.5 % (week of 19 October). The mean week incidence was 0.7%. Overall, we calculated that 20.3 % of the investigated cohort became infected with SARS-CoV-2 in the period from 27 April to 15 November. 3.1 % of Antwerp province inhabitants tested positive in the investigated period (Sciensano data). In August 2020, 1.4 million Belgians (12.2% of Belgian population) filled in a Passenger Locater Form. 2% of travelers tested positive for SARS-CoV-2. On population level, this equals a potential increase of 0.2% in SARS-CoV-2 carrier-rate.

**Figure 2.**
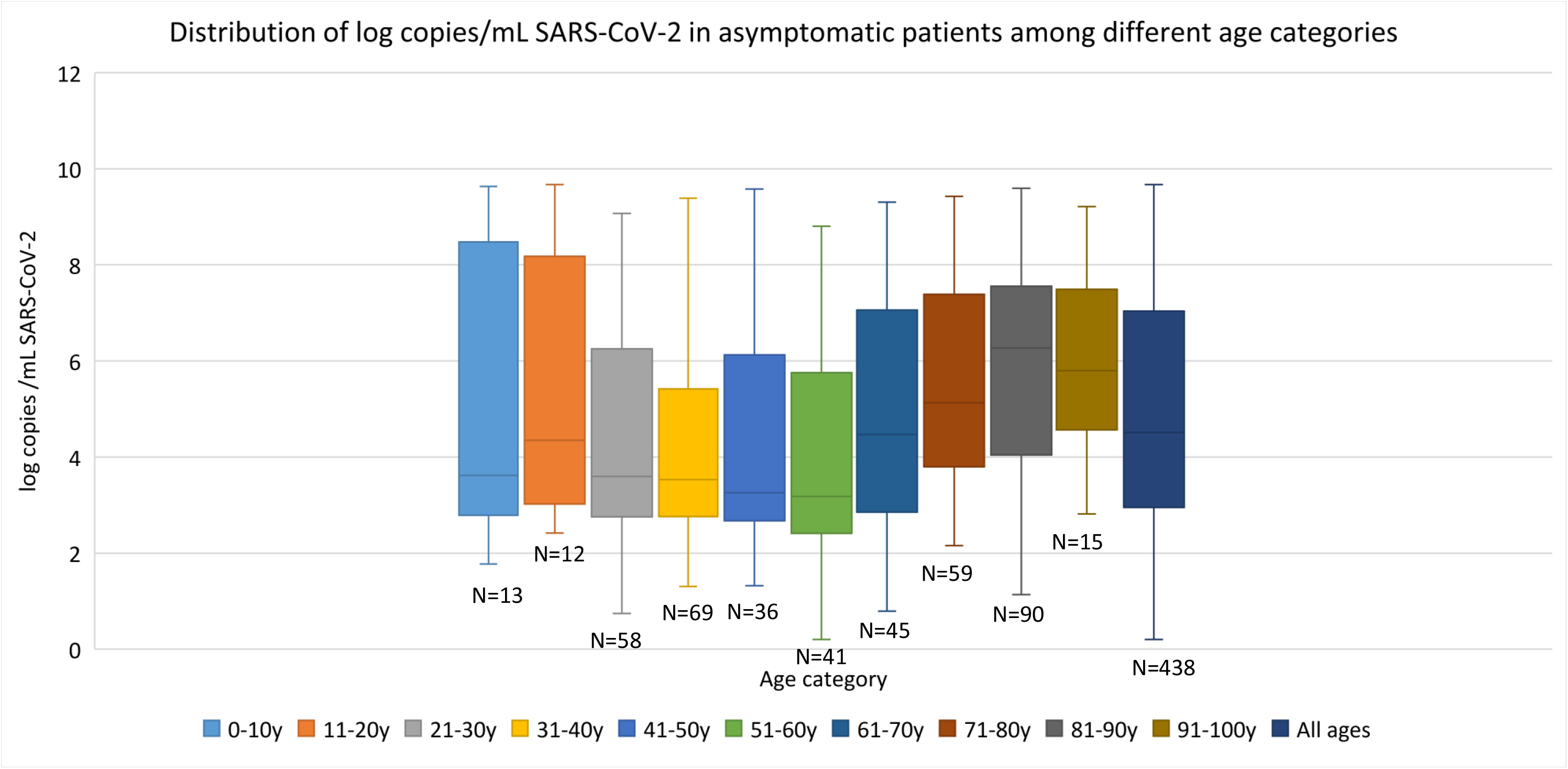
Viral loads according to age groups in asymptomatic carriers (viral loads only available for GZA and ZNA).

The percentage of weekly new hospital admissions varied between 0.00% (see Figure 1) and 0.02% (week of 26 October, 2 November, and 9 November) of the Antwerp province population. The mean ratio of weekly detected cases versus the estimated weekly incidence was 29.1 % (range: 2.9 % - 100 %). The percentage of successfully contacted positive cases varied between 54.0% (week of 22 June) and 94.0 % (week of 26 October) for the Antwerp province with a mean percentage of successfully contacted positive cases of 83.0% (data only available from 1 June to 15 November since registration only started on 1 June).

The SARS-CoV-2 positivity rate according to age is 0.9% (95 % Confidence Interval (CI): 0.4 – 1.5 %) (n= 1292) for daycare children (0-4 years), 1.0% (95 % CI: 0.5 – 1.9 %) (n= 877) for primary school children (5-12 years), 0.8% (95 % CI: 0.3 – 1.6 %) (n = 878) for secondary school children (13-18 years), 0.9% (95 % CI: 0.6 – 1.3 %) (n= 3047) for overall school-aged civilians (0-18 years), 1.4% (95 % CI: 1.3 – 1.5 %) (n = 35637) for adults (>18 years) and 2.6% (95 % CI: 2.1 – 3.2 %) (n = 4033) for the elderly (>80 years). The distribution of viral loads and ratios of carriers with high viral loads among different age categories are presented in Figure 2 and 3 respectively.

**Figure 3.**
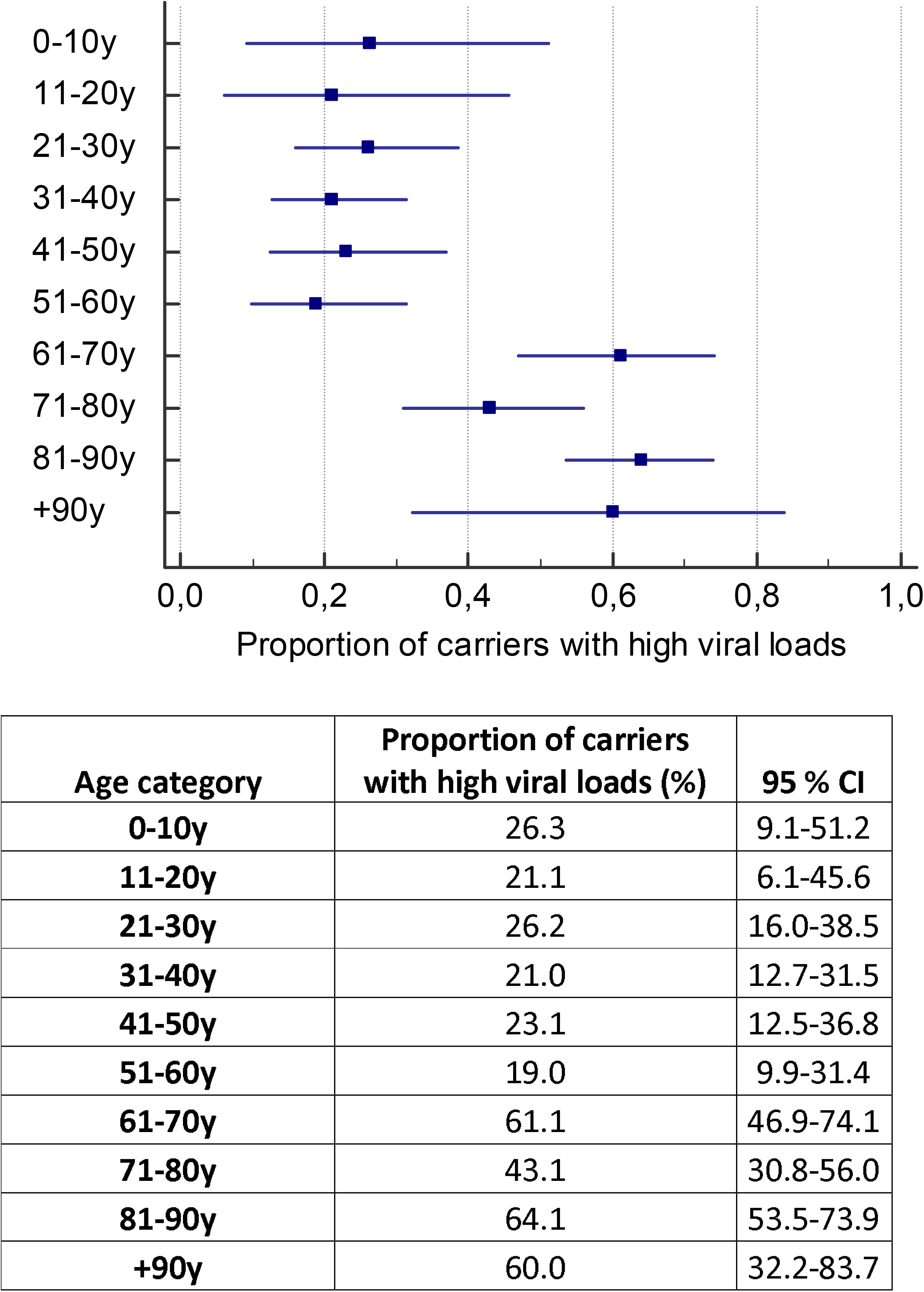
Proportion of asymptomatic carriers with high viral loads according to age category.

The results of the Oxford Stringency Index are shown in Figure 1.

## DISCUSSION

Numerous reports provide data indicating that asymptomatic (or pre-symptomatic) subjects can transmit COVID-19 with high efficiency^10,11^. In the living review paper of Buitrago-Garcia, the secondary attack rate is only slightly lower in contacts of people with asymptomatic infection than in those with symptomatic infection (relative risk 0.35, 95% CI 0.10-1.27)^12^. Mathematical modelling studies (not peer reviewed) have suggested that asymptomatic individuals might be major drivers for the growth of the COVID-19 pandemic^13^.

We found high positivity ratios of asymptomatic carriership in our investigated cohort of Antwerp inhabitants throughout the investigated pandemic period (27 April to 14 November 2020). Lowest positivity ratios were seen at the end of June (week of 29 June): after a period of a full lockdown and only minor easing of containment measures, SARS-CoV-2 positivity ratios were found to be at levels of 1 out of 1000 samples. Similar or even far lower prevalences led to massive screening programs in China in order to stop further viral spread^14^.

Taking into account the SARS-CoV-2 key transmission epidemiological parameters (estimated R_0_ factor of 2.0 and a dispersion factor of 0.10), a strategy accepting prevalences above 1% (the overall average in our cohort being 1.3 %) together with the simultaneously reopening of a locked community were, in our opinion, the perfect ingredients for rapid escalation towards a subsequent second national wave^15^. This finding is important as Belgium has chosen a strategy of accepting significant circulation of the virus within the population without having an accurate insight of past and actual viral prevalences (in contrast to eg. Luxemburg and China, where thorough search and/or isolation programs have been set up)^14,16^. Also, pressure is already increasing to further ease the contingency measures taken in order to control the second wave.

The finding that, due to the implemented testing and tracing strategy, only marginal numbers of positive cases were detected (we estimated that, by extrapolation of cohort data, 20.3 % of the Antwerp population contracted the virus in the study period, but only 3.1 % (15.3% of estimated positieve cases) were effectively tested positively) and subsequently traced (83, range: 54 % - 94 %), even in the periods of upward movement of the curves, was probably another important contributing factor to the uncontrollable rapid further viral spread seen in October 2020. The hypothesis of underestimated viral circulation in Belgium has also been put forward by Herzog and colleagues, by analyzing epi-serological data^17^. Since we only measured the base-line (population wide) prevalence, our estimation may even be an underestimation of the real viral circulation, since outbreaks and clusters are not represented in our analysis, but may have been picked up in documented testing numbers (Sciensano). On the other hand, the underrepresentation of the younger age groups versus the overrepresenation of the older age groups in the investigated cohort leads to an upward bias.

Furthermore, backward tracing has not been implemented optimally to date by the governmental task force^18^. We argue that the Belgian testing and tracing strategy was not able to contain and monitor the pandemic in Belgium adequately, this being one of the major ingredients of a failing containment policy leading to the second national wave.

The province of Antwerp was one of the three Belgian provinces (out of 10) with highest number of tests per inhabitants^4,5^. There is no reason for postulating an exceptional role of Antwerp province in Belgium as being a highly affected province. Inversely, the southern part of the country was affected more severely in the second wave, as shown in the governmental Sciensano data.

Interestingly, we also found higher proportions of carriers in the city of Antwerp (1.8%) than in the province of Antwerp (1.2%; *p* <0.01). This reflects a national trend: the second wave was more prevalent in crowded bigger capital cities^4^.

With regard to positivity ratios according to age category, we found high numbers (around 1 % throughout the investigated period) in school children and young adults (0-30 years). Positivity ratios of young adults (21-40 years-old) were significantly higher than the adult population aged 51-70 years (*p* <0.01).

Although the role of children as main drivers of transmission is still debated, our finding may be another argument for active transmission in these age groups^20,21,22^. A recent seroprevalence survey in two Belgian municipalities (one highly affected, the other hardly affected) also showed equivalent proportions of infections in primary school children versus adolescents in the first three years of secondary education^23^.

On the other hand, if children were equally susceptible, one might expect even higher rates of carriership, since children were less restricted in the number of contacts (summercamps, schools were open from 1 September 2020). Since we were not able to correct for number of contacts, we cannot further determine the value of this hypothesis.

With regard to age distribution, another interesting finding is the high overall percentage of positivity ratios in the elderly (+70 y). In our hypothesis, this is the reflection of the augmented transmission risks in residential care homes.

We also found significant proportions of carriers with high viral loads (on average 35.4 % of all inclusions). In the week of 26 October, we estimate that 7.0% of Antwerp inhabitants were carrying high viral loads. Evaluating the whole investigated period, on average 0.7% of the population was carrying high viral loads. Although the link of viral load with contagiousness has not been established, this might have been an important factor in the spread of the virus^19^.

Viral loads in younger age groups were not lower than viral loads in the working age population. (Figure 2) In another non-peer-reviewed publication, it was also shown that there is no significant difference between viral loads in 1-20 year-olds as compared to 21-100 year-old adults^24^. Furthermore, another study suggests that the viral load in children below 5 years of age with mild to moderate COVID-19 symptoms is higher than in older children and adults^25^.

In our hypothesis, positive children may have further introduced the virus into households, which suggests that this is a potentially underestimated factor in the derailing of the situation in September 2020.

Interestingly, we found significantly higher proportions of carriers with high viral loads in the elder age categories. The combination of higher prevalences and higher ratios of carriers with high viral loads in this age group might be an important driver of the abundance of outbreaks and associated excess morbidity and mortality seen in retirement homes.

Overall, not recognizing the presence and potential important role of the asymptomatic carriers with higher viral loads may also have been another ingredient which led to the subsequent second wave.

In our opinion, the role of the returning travelers was very limited. In August 12.2 % of the Belgian population filled in a Passenger Locater form. About 2% of returning travelers tested positive. On a population level, this may have led to an increase of 0.2% in carriership rates.

On the positive side, we found that by implementing strict contingency measures (i.e. following the tightening and easing advice of experts) even substantial viral circulation of around 1% can be controlled: the ratio of positive carriers was contained in the period between 27 April and 14 September (19 weeks) without overstretching health care capacities. However, the efforts required to maintain controllable containment were enormous, thereby putting high pressure on social (limited number of contacts) and economic well-being (Oxford Stringency Index not below 50.0 between 27 April and 14 September).

Finally, we come to our last driver of the second wave, probably the most important one. Mid to end September, key Belgian epidemiologists, virologists and biostatisticians warned the temporary government of the possible rapid escalation in numbers of new infections and admissions, based on available data. The government decided not to follow their advice, and implemented a strategy that eased restriction measures (Oxford Stringency Index on 14 September: 52.8; 5 October: 47.2) in a time when there was a steadily increasing number of new infections (incidence increased from 0.9% the week of 14 September to 2.3% the week of 5 October). They were being encouraged to carry out this strategy by various pressure groups. In October, Belgium was facing a second wave with a rapid increase in numbers of COVID admissions, thereby highly impacting the health care system and hospital facilities.

The limitations of this study include that (weak positive) asymptomatic cases could have corrresponded to previous infections with persistant existence of detectable RNA. This may have biased the results. However, the rapid decrease of positivity ratios after the implementation of extra measures (second pandemic wave) suggests we predominantly included active infections. Although the phenomenon of prolonged shedders does certainly exist, our data suggest that it is relatively insignificant and does not affect the major findings in this paper. Also, the median duration of positive results has been shown to be two to three weeks^4^. Another limitation is that we did not follow up positive/negative cases. We did not link the data to secondary COVID-19 cases, thus not establishing the contagiousness of positive cases. The screenings were linked to hospital procedures/admissions. Although we can accept these screenings as random samples from the community, the cohort is not a representative sample from the community: data points were not equally distributed in the Antwerp province, and age distributions were not representative for the Antwerp province (underestimation of younger age groups, and overestimation of older age groups).

In conclusion, we found that SARS-CoV-2 was significantly present in the Antwerp province population in the period after the first wave. The virus circulated in all age groups with a predomincance in the elderly. 1.3% of the investigated cohort tested positive for SARS-CoV-2 with 35% carrying high viral loads. The testing and contact-tracing policy only detected a small proportion of the real viral circulation. The restriction measures were eased in a period of increasing incidences. The combination of these factors were, in our opinion, the ingredients for the onset of the second wave. If Belgium chooses to accept a certain viral circulation, a very strict barometer has to be put into place, in order to prevent further worsening of the pandemic situation. The positivity rates in (pre)admission screening results may be an important tool;

**Table 1.** Age distribution of included cases and age distribution of the province of Antwerp.

| **Age category** | **Number of inclusions** | **Percentage of inclusions** | **Percentage of inhabitants in the Antwerp province** | ***p*-value (difference in rates of inclusions versus the Antwerp province)** |
| --- | --- | --- | --- | --- |
| **0-10 y** | 1971 | 5.1 | 11.3 | *p*<0.01 |
| **11-20 y** | 1493 | 3.9 | 11.0 | *p*<0.01 |
| **21-30 y** | 3942 | 10.2 | 12.1 | *p*<0.01 |
| **31-40 y** | 5110 | 13.2 | 13.1 | *p*=0.83 |
| **41-50 y** | 4376 | 11.3 | 12.7 | *p*<0.01 |
| **51-60 y** | 5993 | 15.5 | 14.0 | *p*<0.01 |
| **61-70 y** | 6194 | 16.0 | 11.8 | *p*<0.01 |
| **71-80 y** | 5651 | 14.6 | 8.2 | *p*<0.01 |
| **81-90 y** | 3450 | 8.9 | 4.9 | *p*<0.01 |
| **+90 y** | 583 | 1.5 | 1.0 | *p*=0.23 |

**Table 2.** Overall positivity rate of SARS-CoV-2 in asymptomatic carriers according to age groups.

| Age category | Overall percentage of positive tests | 95% CI |
| --- | --- | --- |
| **0 - 10 y** | 1.0 | 0.6 to 1.5 |
| **11 - 20 y** | 1.3 | 0.8 to 2.0 |
| **21 - 30 y** | 1.6 | 1.3 to 2.1 |
| **31 - 40 y** | 1.6 | 1.3 to 2.0 |
| **41 - 50 y** | 1.2 | 0.9 to 1.6 |
| **51 - 60 y** | 1.0 | 0.7 to 1.2 |
| **61 - 70 y** | 0.9 | 0.7 to 1.1 |
| **71 - 80 y** | 1.2 | 0.9 to 1.5 |
| **81 - 90 y** | 2.7 | 2.2 to 3.3 |
| **> 90 y** | 2.6 | 1.4 to 4.2 |

## Data Availability

Data are available upon request.

## Acknowledgements

None

## Author Bio

Dr. Reinout Naesens is Head of the Infection Prevention and Control Department and member of the Medical Microbiology Department of the ZNA group, the largest public hospital in Belgium.

## REFERENCES

1. World Health Organisation, World Health Organisation situation reports, https://www.who.int/emergencies/diseases/novel-coronavirus-2019/situation-reports. Accessed 2 december 2020.

2. Byambasuren O, Cardona M, Bell K, Clark J, McLaws M-L, Glasziou P. Estimating the extent of true asymptomatic COVID-19 and its potential for community transmission: systematic review and meta-analysis. medRxiv. 2020; (published online September 13.) (preprint) doi: https://doi.org/10.1101/2020.05.10.20097543

3. Castagnoli R, Votto M, Licari A, Brambilla I, Bruno R, Perlini S, et al. Severe Acute Respiratory Syndrome Coronavirus 2 (SARS-CoV-2) Infection in Children and Adolescents: A Systematic Review. JAMA Pediatr. 2020;174:882–889. doi:10.1001/jamapediatrics.2020.1467.

4. Sciensano, https://epistat.wiv-isp.be/covid/. Accessed 2 december 2020.

5. Toezicht Volksgezondheid, https://zorgatlas.vlaanderen.be. Accessed 2 december 2020.

6. Statistiek Vlaanderen, https://www.statistiekvlaanderen.be/nl/bevolking-naar-leeftijd-en-geslacht. Accessed 2 december 2020.

7. Oxford COVID-19 Government Response Tracker, https://covidtracker.bsg.ox.ac.uk/. Accessed 2 december 2020.

8. Cevik M, Tate M, Lloyd O, Maraolo AE. SARS-CoV-2, SARS-CoV, and MERS-CoV viral load dynamics, duration of viral shedding, and infectiousness: a sytematic review and meta-analysis. Lancet Microb. 2020. Online first. doi: https://doi.org/10.1016/S2666-5247(20)30172-5.

9. Bullard J, Dust K, Funk D, Strong JE, Alexander D, Garnett L, et al. Predicting infectious SARS-CoV-2 from diagnostic samples. Clin Infect Dis. 2020;22:ciaa638. doi: 10.1093/cid/ciaa638.

10. Koh WC, Naing L, Chaw L, Rosledzana MA, Alikhan MF, Jamaludin SA, et al. What do we know about SARS-CoV-2 transmission? A systematic review and meta-analysis of the secondary attack rate and associated risk factors. PLoS One. 2020;15(10):e0240205. doi: 10.1371/journal.pone.0240205.

11. Mizumoto K, Kagaya K, Zarebski A, Chowell G. Estimating the asymptomatic proportion of coronavirus disease 2019 (COVID-19) cases on board the Diamond Princess cruise ship, Yokohama, Japan, 2020. Euro Surveill. 2020;25(10):2000180. doi: 10.2807/1560-7917.ES.2020.25.10.2000180.

12. Buitgrago-Garcia DC, Egli-Gany D, Counotte MJ, Hossmnn S., Imeri H, Ipekci AM, et al. Asymptomatic SARS-CoV-2 infections: a living systematic review and meta-analysis. medRxiv. 2020; (published online July 28.) (preprint) doi: https://doi.org/10.1101/2020.04.25.20079103.

13. Dobrovolny HM. Modeling the role of asymptomatics in infection spread with application to SARS-CoV-2. PLoS One 2020; 15:e0236976. doi: 10.1371/journal.pone.0236976.

14. Health Commission of Hubei Province. Daily report on epidemic situation of COVID-19 in Hubei province. http://wjw.hubei.gov.cn/bmdt/ztzl/fkxxgzbdgrfyyq/xxfb/index.shtml. Accessed 2 december 2020.

15. Kucharski AJ, Russell TW, Diamond C, Liu Y, Edmunds J, Funk S, Eggo RM; Centre for Mathematical Modelling of Infectious Diseases COVID-19 working group. Early dynamics of transmission and control of COVID-19: a mathematical modelling study. Lancet Infect Dis. 2020; pii:S1473-3099(20)30144-4.

16. https://covid19.public.lu/en/testing.html Accessed 2 december 2020.

17. Herzog S, Abrams S, Wouters I, Ekinci E, Pateet L, Coppens A, et al. Seroprevalence of IgG antibodies against SARS coronavirus 2 in Belgium – a serial prospective crosssectional nationwide study of residual samples. medRxiv. 2020; (published online October 1.) (preprint) doi: https://doi.org/10.1101/2020.06.08.20125179.

18. Endo A; Leclerc QJ, Knight GM, Medley GF, Atkins KE, Funk S, et al. Implication of backward contact tracing in the presence of overdispersed transmission in COVID-19 outbreaks. Wellcome Open Res. 2020;5:239. doi: 10.12688/wellcomeopenres.16344.1.

19. Zou L, Ruan F, Huang M, Liang L, Huang H, Hong Z, et al. SARSCoV-2 Viral Load in Upper Respiratory Specimens of Infected Patients. N Engl J Med. 2020;382:1177-1179. doi: https://www.nejm.org/doi/full/10.1056/NEJMc2001737.

20. Jones TC, Mühlemann B, Veith T, Biele G, Zuchowski M, Hoffmann J, et al. An analysis of SARS-CoV-2 viral load by patient age. medRxiv. 2020; (preprint) doi: https://doi.org/10.1101/2020.06.08.20125484.

21. Viner RM, Russell SJ, Croker H, Packer J, Ward J, Stansfield C, et al. School closure and management practices during coronavirus outbreaks including COVID-19: a rapid systematic review. Lancet Child & Adolescent Health. 2020;4(5):397–404. doi: 10.1016/S2352-4642(20)30095-X.

22. Heavey L, Casey G, Kelly C, Kelly D, McDarby G. No evidence of secondary transmission of COVID-19 from children attending school in Ireland, 2020. Euro Surveillance: bulletin Europeen sur les maladies transmissibles = European communicable disease bulletin. 2020;25(21). doi: 10.2807/1560-7917.ES.2020.25.21.2000903.

23. Sciensano, https://www.sciensano.be/sites/default/files/limburg-validation-sars-cov2_report_20201112_final.pdf. Accessed 2 December 2020.

24. Jones TC, Mühlemann B, Veith T, Zuchowski M, Hofmann J, Stein A, et al. An analysis of SARS-CoV-2 viral load by patient age. Available from: https://zoonosen.charite.de/fileadmin/user_upload/microsites/m_cc05/virologie-ccm/dateien_upload/Weitere_Dateien/analysis-of-SARS-CoV-2-viral-load-by-patient-age.pdf. Accessed 2 December 2020.

25. Heald-Sargent T, Muller WJ, Zheng X, Rippe J, Patel AB, Kociolek LK. Age-Related Differences in Nasopharyngeal Severe Acute Respiratory Syndrome Coronavirus 2 (SARS-CoV-2) Levels in Patients With Mild to Moderate Coronavirus Disease 2019 (COVID-19). JAMA Pediatr. 2020;174(9):902–903. doi : 10.1001/jamapediatrics.2020.3651.

